## Supplementary material for "The impacts of COVID-19 mitigation on dengue virus transmission: a modelling study"

### Supplementary Text

Our agent-based model (ABM) includes both people and mosquitoes as agents. These agents interact with each other in an environment represented by a set of locations, including houses, schools, parks, cemeteries, and churches. This environment also incorporates temporally varying climatic conditions, which affect mosquito biting frequency, survival, and extrinsic incubation period for dengue virus (DENV). The environment is based on the city of Iquitos in Peru, and we represent all 92,891 buildings in the city. We use exact spatial coordinates and location type for the 38,835 locations for which these data were available. For locations without these data, we randomly distributed the locations and assigned a location type so that they were evenly spaced and representative of the location types we had data on.

We modeled approximately 450,000 humans in our model, chosen to reflect the demography of Iquitos and its surroundings (1). We based the overall age and sex distribution on U.N. estimates of these for Peru, and based the demographic profile of each household on survey data from a prior study (2). This synthetic population also realistically captured how people are distributed across houses and how demographics changed over time. We achieved this by simulating human births and deaths that match those estimated by the U.N. for Iquitos and simultaneously preserved realistic household compositions by placing newborn children in houses with appropriately aged mothers as determined by U.N. estimates of age-specific fertility of Peru (1). All human agents in the model went through daily human movement patterns using a model previously described by Perkins et al. (3), except when this was modified by lockdown policies which meant they stayed home (see Methods). To describe these movement patterns, each agent has five daily movement trajectories. At the start of each day one of these five trajectories is chosen, with equal probability, for each agent. The human movement patterns were fitted to data from retrospective, semi-structured interviews with inhabitants of Iquitos (4,5).

Immature mosquitoes were modeled deterministically and independently at each location. They transitioned through three immature stages: eggs, larvae, and pupae, with the number of pupae in a house determining the rate of emergence of adult mosquitoes in that house. The rate of transition between each of these stages was temperature dependent. All stages also underwent temperature-dependent mortality. Larval stages underwent an additional density-dependent mortality (6,7). Both the larval and pupal stages also underwent an additional rate of mortality that was calibrated so that adult abundance matched a statistical estimate of the spatio-temporal adult abundance in Iquitos (8). Adult mosquitoes are modeled as agents, and take blood-meals upon co-located human agents. When a mosquito takes a blood-meal, the time of its next blood-meal is determined according to an exponential distribution with a time-varying rate parameter, based on temporal trends in temperature and empirical relationships between these rates and temperature (7,9). When the mosquito's next blood-meal is due, it will take it unless there is no human present. This means that the number of blood-meals taken by a mosquito is not determined by the local density of humans, except in the unusual instance in which no human is present at a location. The mosquito determines which human it will bite as a function of the body sizes of humans present at that time (2). Each day, each mosquito moves to another location with probability 0.3, and will only move to a location within 100 m of its starting location, consistent with another agent-based model of *Ae. aegypti* population dynamics (6).

When a mosquito takes a blood-meal on a human and one of them is infected, transmission can occur. The probability that transmission occurs from humans to mosquitoes is determined by the time since the human was infected, with this probability based on the viremia levels of the infecting human at the time of the bite (10). For transmission from mosquitoes to humans, infectious mosquitoes transmitted DENV to susceptible humans with a fixed probability of 1.0, providing the extrinsic incubation period has been completed (11,12). Once infected humans develop symptoms following a latent period derived from timing of peak viraemia (11). Following recovery, humans become permanently immune to that serotype and also have temporary heterologous immunity to all other serotypes. This temporary immunity lasts for an exponentially distributed period with mean of 686 days, estimated in a previous modeling study of time-varying serotype-specific dengue incidence (14). The initial level of immunity in the model was calibrated to an analysis of longitudinal serological data (15).

### Supplementary Figures

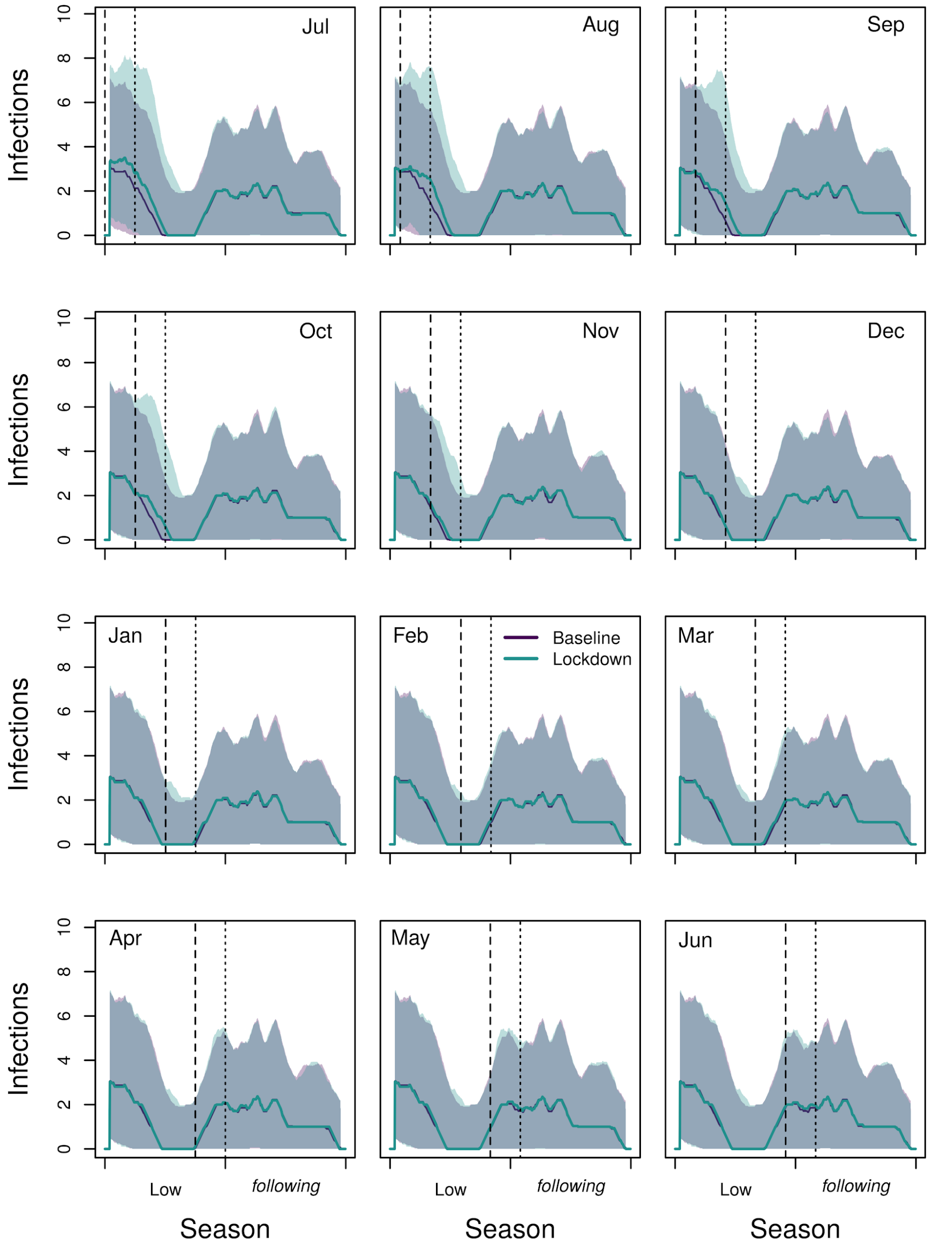


*Figure S1: Time series of local DENV infections across the low-transmission and following seasons when lockdown was initiated on the first of the month (dashed line) in the low season and lasted for two months (ending at the dotted line). Shaded regions are the interquartile range. Shading in gray is where these regions overlap.*

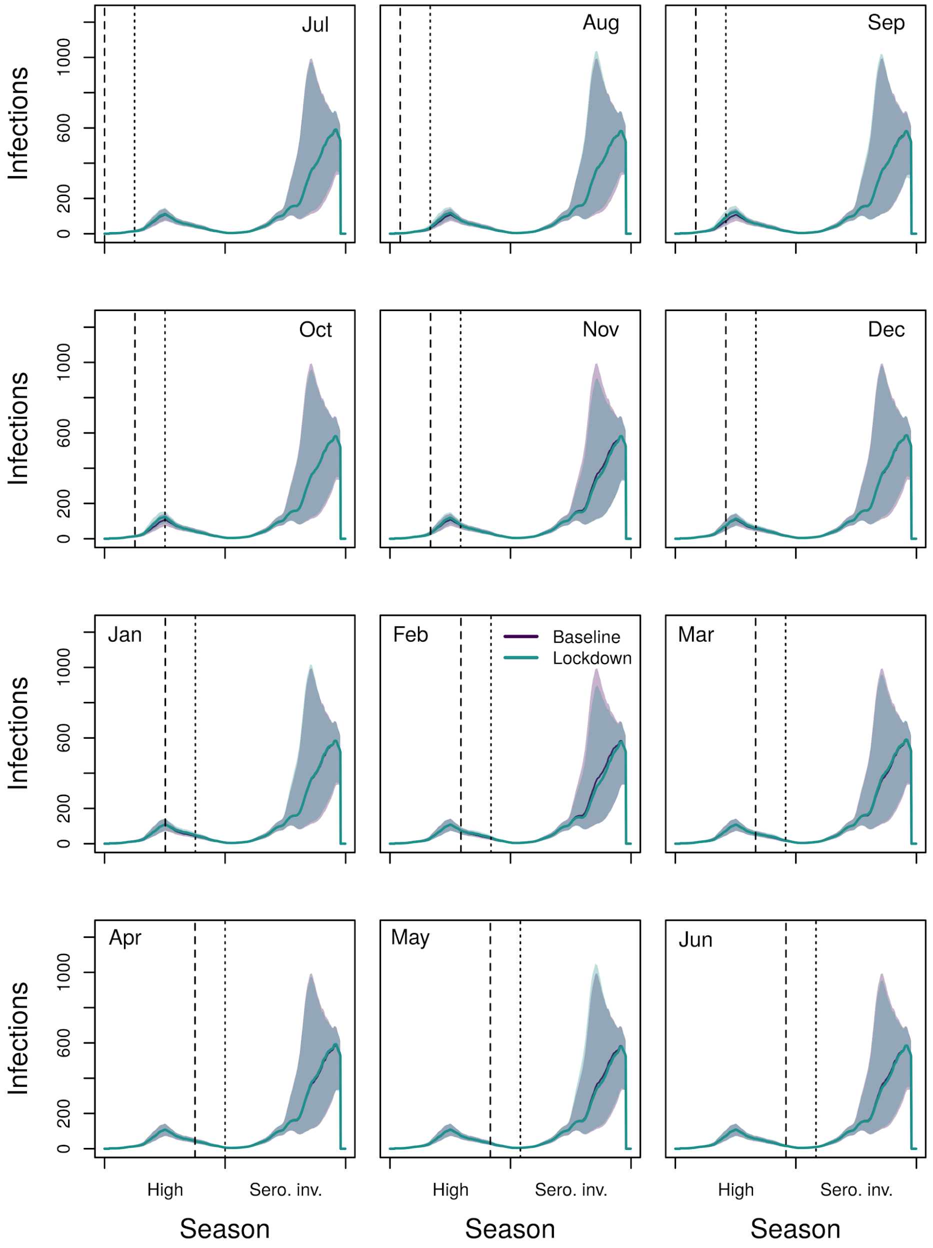


*Figure S2: Time series of local DENV infections across the high and serotype invasion seasons when lockdown was initiated on the first of the month (dashed line) in the high season and lasts for two months (ending at the dotted line). Shaded regions are the interquartile range. Shading in gray is where these regions overlap.*

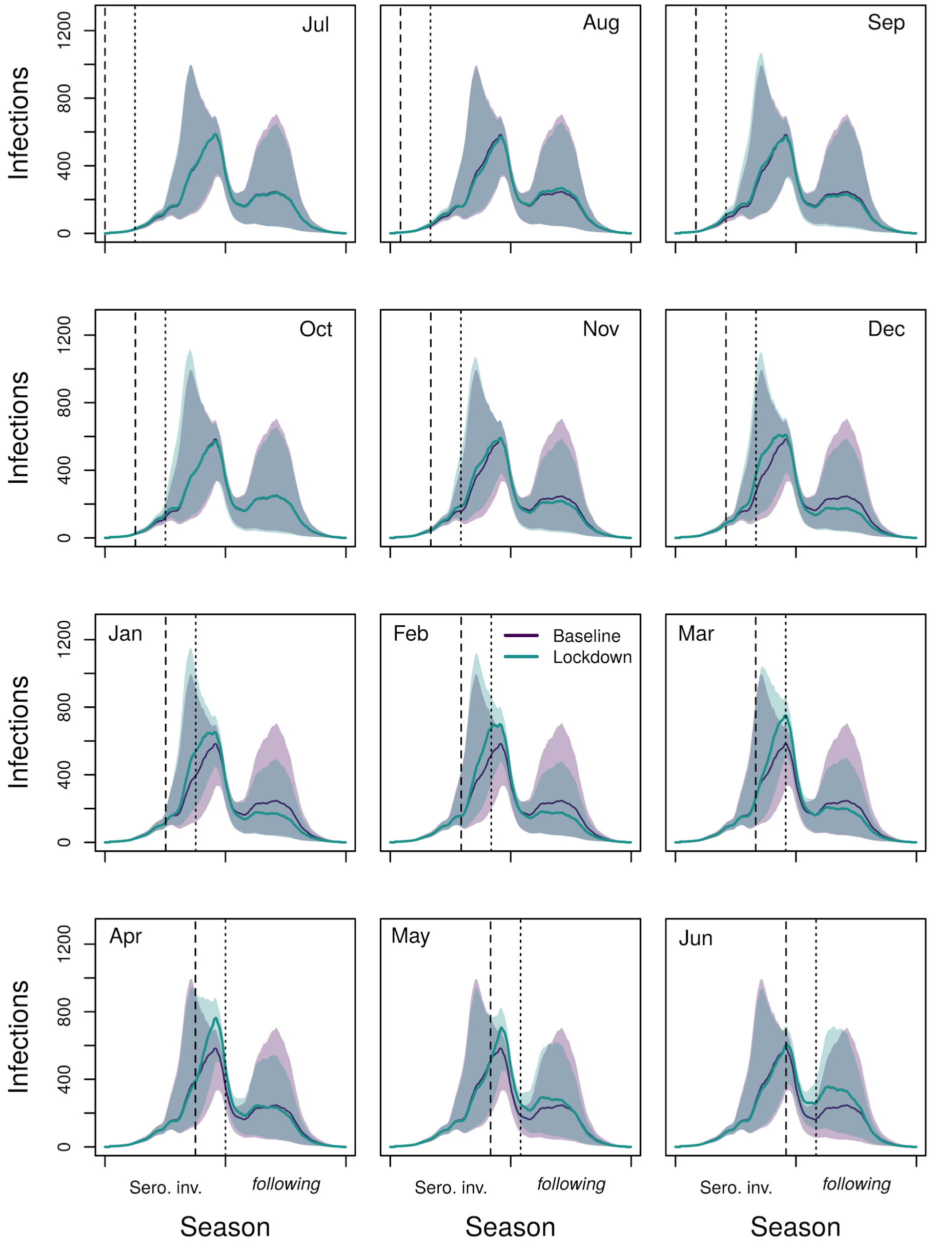


*Figure S3: Time series of local DENV infections across the serotype invasion and following seasons when lockdown was initiated on the first of the month (dashed line) in the* serotype invasion *season and lasts for two months (ending at the dotted line). Shaded regions are the interquartile range. Shading in gray is where these regions overlap.*

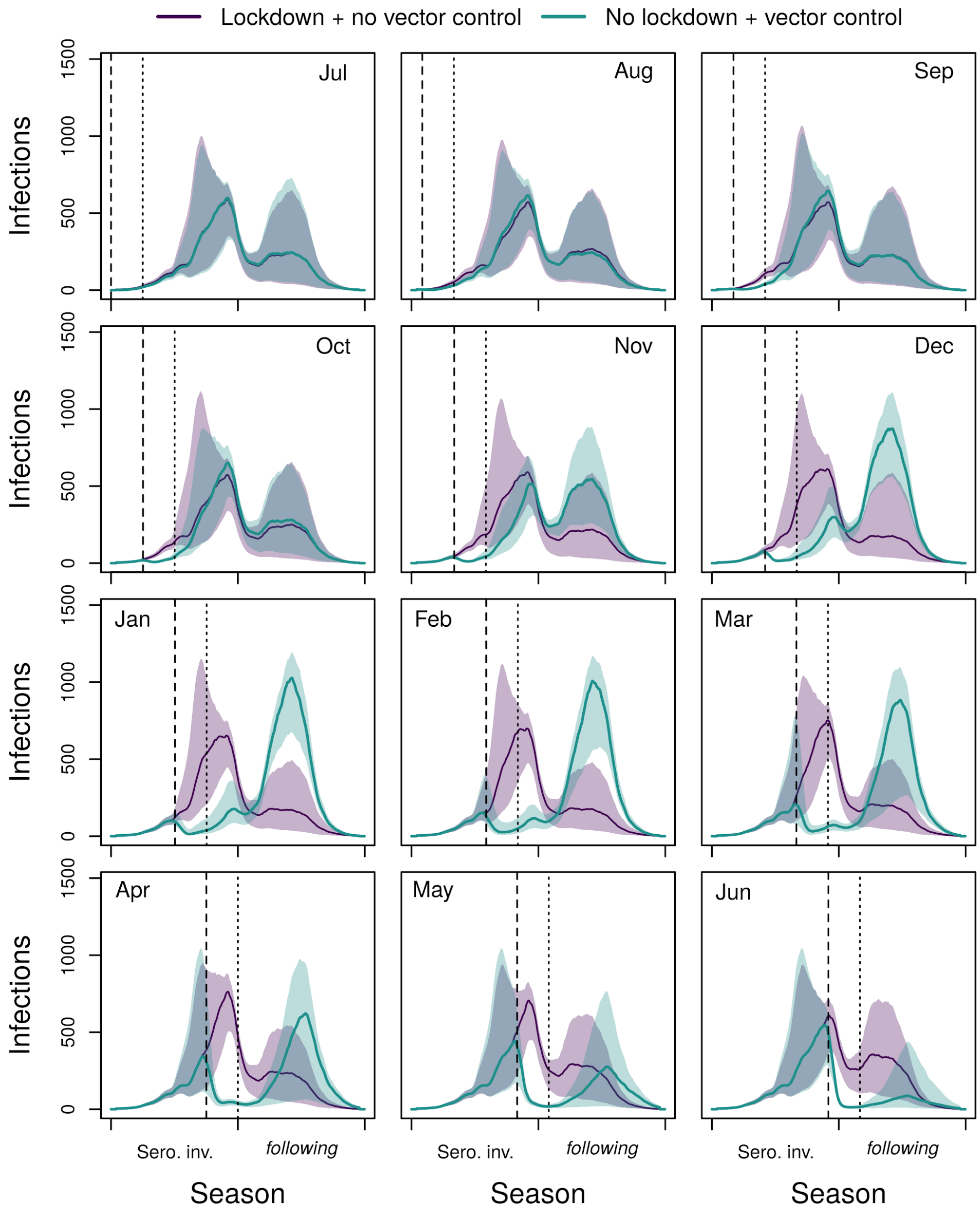


*Figure S4: Time series of local DENV infections in the serotype invasion scenario, comparing lockdown without vector control (purple) to no lockdown with vector control (green). Lockdown and the city-wide vector control campaign began at the dashed line. Lockdown lasted three months (ending at the dotted line). The vector control campaign lasted three weeks. Shaded regions are the interquartile range. Shading in gray is where these regions overlap.*

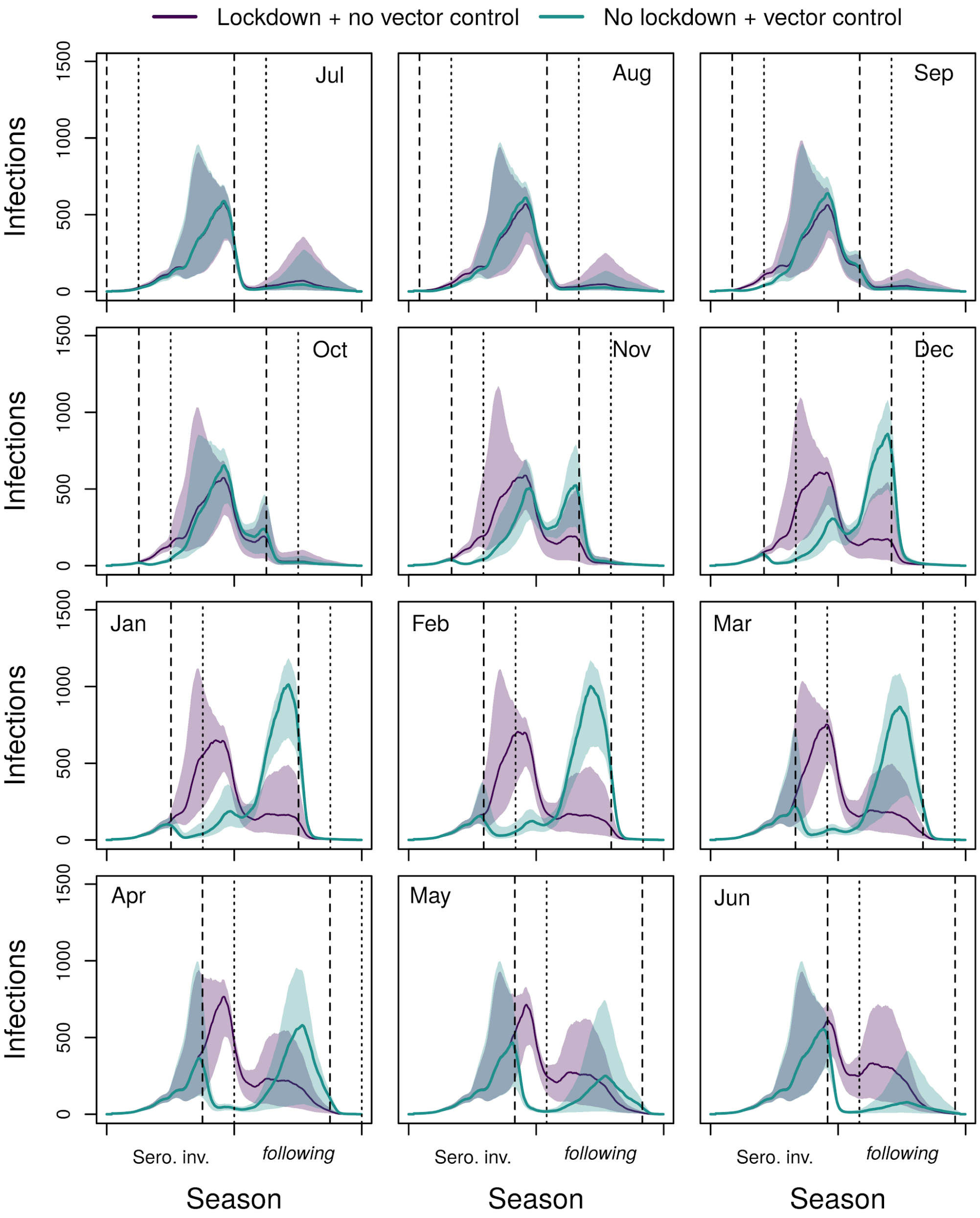


*Figure S5: Time series of local DENV infections in the serotype invasion scenario, comparing lockdown without vector control (purple) to no lockdown with vector control (green). Both lockdown and the city-wide vector control campaign began at the dashed line in the first season. In the following season, vector control occurred in both simulations and lockdown did not occur in either simulation. Lockdown lasted three months (ending at the dotted line). The vector control campaign lasted three weeks. Shaded regions are the interquartile range. Shading in gray is where these regions overlap.*

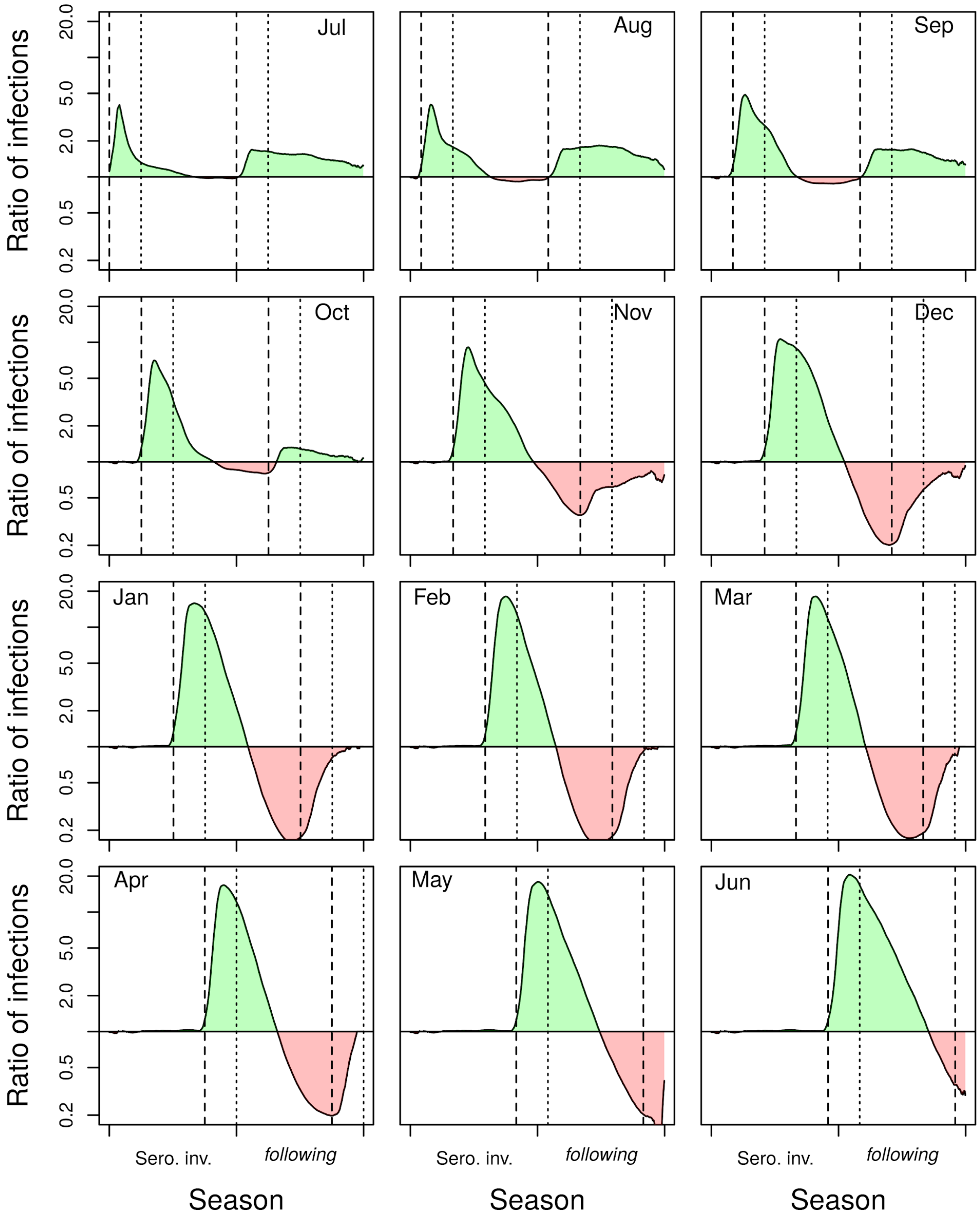


*Figure S6: Ratio of the mean number of infections under lockdown to the mean number in the baseline scenario without lockdown in the serotype invasion and following season. Lockdown began at the vertical dashed line, and ended at the dotted line. In the following season, vector control occurred in both simulations and lockdown did not occur in either simulation.*

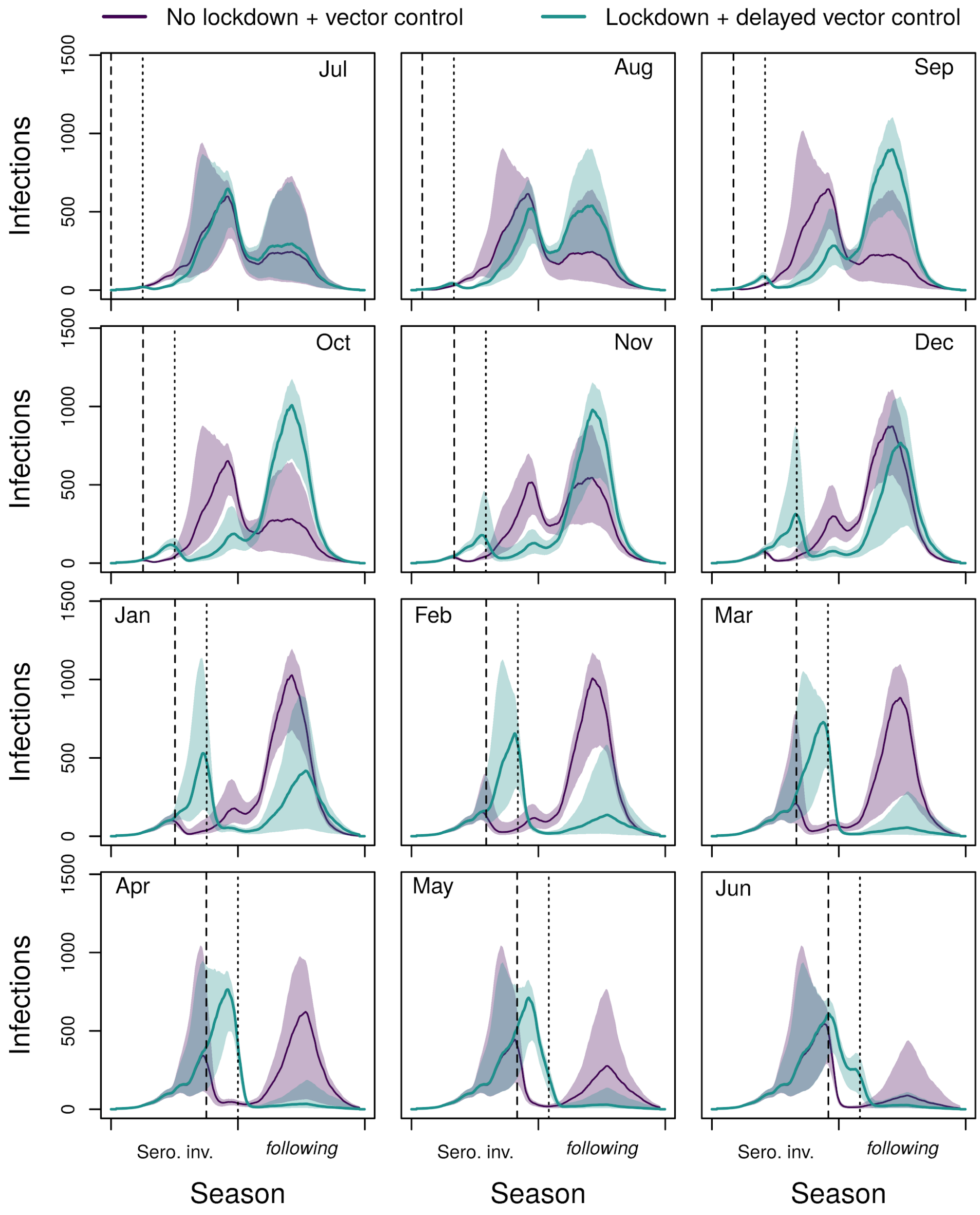


*Figure S7: Time series of local dengue infections in the serotype invasion scenario, comparing vector control without lockdown (purple) to lockdown with a vector control campaign which begins as soon as lockdown ends (green). Lockdown lasted three months, starting at the dashed line and ending at the dotted line. The vector control campaign lasted three weeks, beginning at the dashed line (purple) or the dotted line (green). Shaded regions are the interquartile range. Shading in gray is where these regions overlap.*

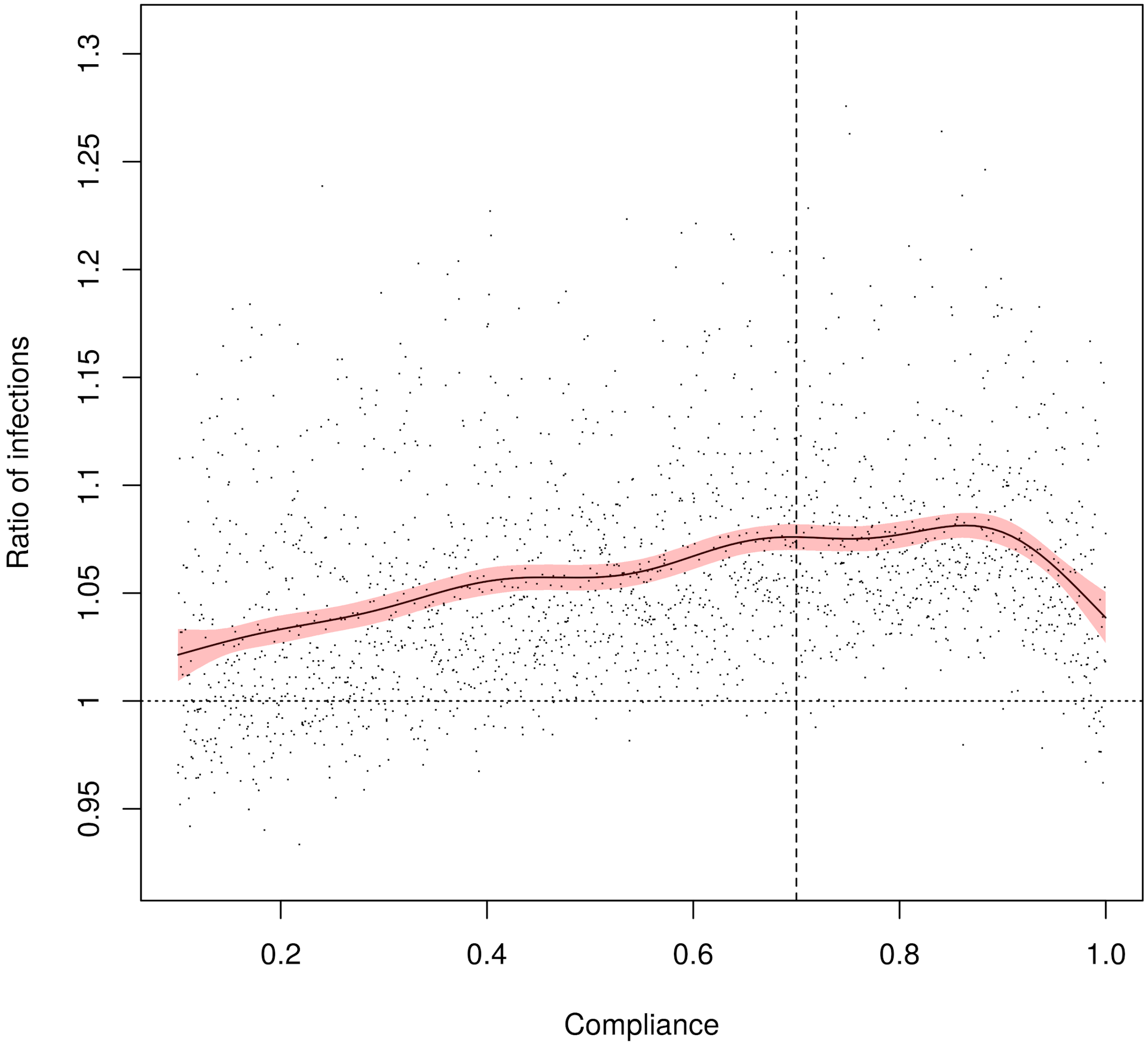


*Figure S8: Effect of changing lockdown compliance on the proportional change in the cumulative number of infections across two consecutive seasons. The vertical dashed line shows baseline compliance (70%). The horizontal dashed line shows when there is no effect of lockdown.*

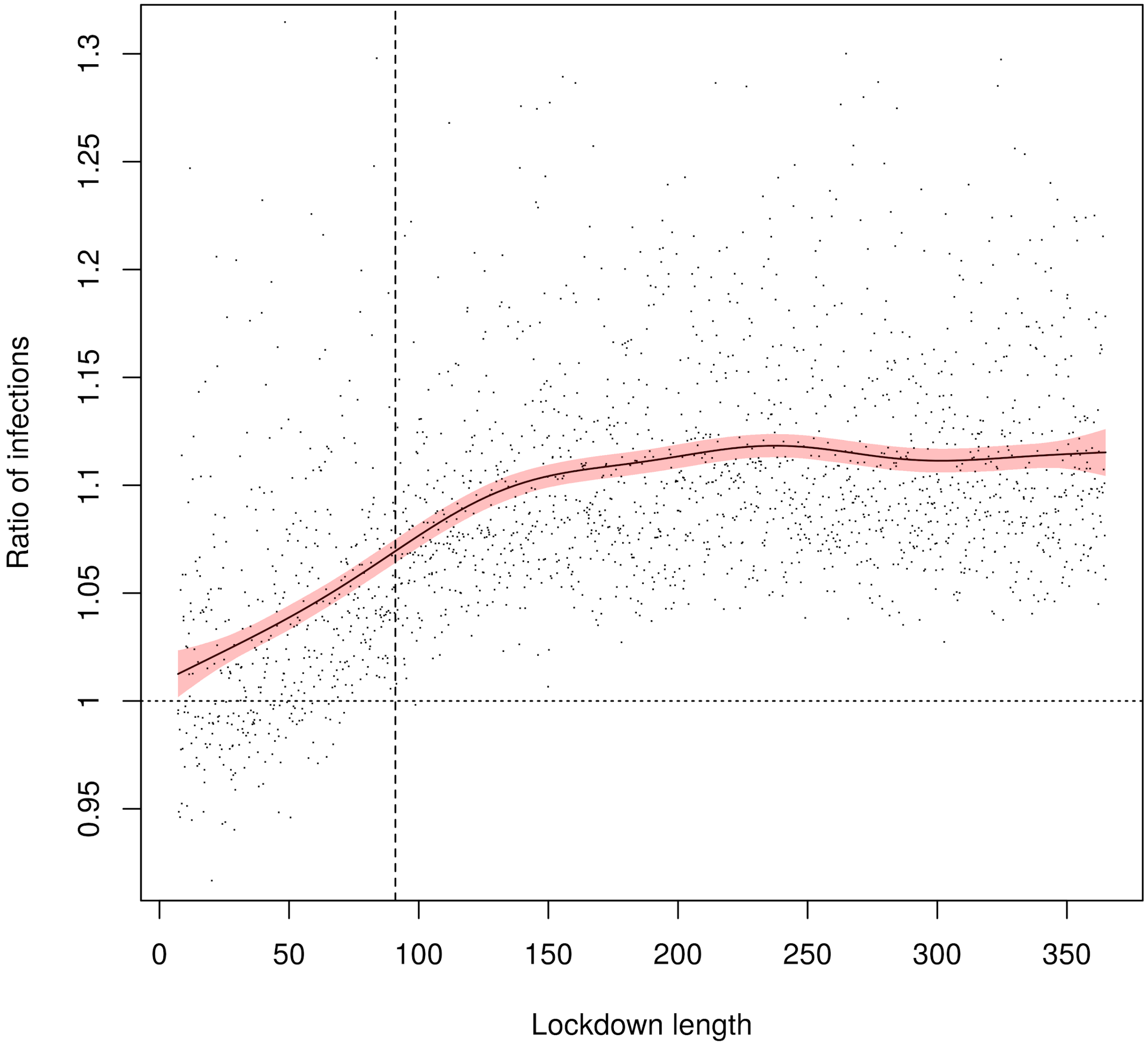


*Figure S9: Effect of changing lockdown length on the proportional change in the cumulative number of infections across two seasons. The vertical dashed line shows the baseline length (three months). The horizontal dashed line shows when there is no effect of lockdown.*
